# Missing the vulnerable – Inequalities in social protection among the general population, people living with HIV, and adolescent girls and young women in 13 sub-Saharan African countries: Analysis of population-based surveys

**DOI:** 10.1101/2024.02.08.24302524

**Authors:** David Chipanta, Silas Amo-Agyei, Lucas Hertzog, Ahmad Reza Hosseinpoor, Michael Smith, Caitlin Mahoney, Juan Gonzalo Jaramillo Meija, Olivia Keiser, Janne Estill

## Abstract

Inequality in access to services is a global problem mainly impacting the poorest populations. The role of social protection in reducing inequalities is recognized, but few studies have investigated whether social protection benefits people facing considerable socioeconomic inequalities. We assessed inequalities in receiving social protection among the public, men and women living with human immunodeficiency virus (PLHIV), and adolescent girls and young women (AGYW), using population-based data from 13 African countries. We constructed concentration curves and computed concentration indices (CIX) for each country and population group. We also conducted a desk review of social protection in the studied countries where information was available on the characteristics of social protection programs and their access by the general population, PLHIV, and AGYW. The sample size ranged from 10,197 in Eswatini to 29,577 in Tanzania. Women comprised 60% or more of PLHIV in the surveyed countries. 50%–70% of the respondents were unemployed, except in Cameroon, Kenya, and Uganda, where less than 50% were unemployed. Generally, the proportion of respondents from wealth quintile one (Q1), the poorest 20% of households, was like that from Q2–Q5. The proportion of the general population receiving social protection varied from 5.2% (95% Confidence Interval 4.5%– 6.0%) in Ethiopia to 39.9% (37.0%–42.8%) in Eswatini. Among PLHIV, the proportion receiving social protection varied from 6.9% (5.7%–8.4%) among men living with HIV in Zambia to 45.0% (41.2–49.0) among women living with HIV in Namibia. Among AGYW, the proportion varied from 4.4% (3.6–5.3) in Ethiopia to 44.6% (40.8–48.5) in Eswatini. In general, 15% or less of the respondents from Q1 reported receiving social protection in eight countries (i.e., Cameroon, Côte d’Ivoire, Ethiopia, Kenya, Malawi, Tanzania, Uganda, and Zambia), with 10% or less in three countries (Cameroon, Côte d’Ivoire, and Ethiopia); 15%– 20% in Rwanda, 30% in Zimbabwe, 40% in Lesotho, and more than 50% in Eswatini and Namibia. Among the wealthiest quintiles (Q5), the proportion receiving social protection ranged from 3.6% (2.6%–5.0%) in Ethiopia to 19.7% (16.25–23.8%) in Namibia. Only in countries with higher social protection coverage did the proportion of the poorest wealth quintile households reached also high. Socioeconomic inequalities in receiving social protection favored the poor in 11 out of 13 countries and the rich in Cameroon and were undefined in Côte d’Ivoire. The CIX values for socioeconomic inequalities in receiving social protection in these 11 countries ranged from −0.080 (p=0.002) among the general population in Malawi to −0.372 (p< 0.001) among WLHIV in Zimbabwe. However, in 8 countries (Cameroon, Côte d’Ivoire, Ethiopia, Kenya, Malawi, Tanzania, Uganda, and Zambia) of these 11 countries, 15% or less of the population from the poorest wealth quintile received social protection. In the countries surveyed, access to social protection for the general population, MLHIV and WLHIV, and AGYW was generally low but favored people from poor households. However, pro-poor social protection, although necessary, is not sufficient to ensure that people from the poorest households receive social protection. Further research is required to identify and reach people from the poorest households with social protection in sub-Saharan Africa.

## 1 Introduction

Inequality in access to services is an urgent global concern, with the alarming gap between extreme wealth and poverty reaching unprecedented levels and thrusting billions of people into hardship, including hunger [1, 2]. Inequality is multifaceted, spanning race, ethnicity, income, wealth, and gender. Gender inequalities are deeply entrenched and intersect with other forms of inequality [3, 4]. Generally, equitable communities enjoy robust social cohesion, low crime rates, high levels of trust, life satisfaction, durable peace, political stability, and economic growth, in contrast to their inequitable counterparts [3, 5, 6, 7]. Conversely, high inequalities can undermine a nation’s capacity to prevent, respond, and adapt to emergencies, including infectious diseases [3, 7]. Therefore, addressing inequalities is an imperative objective of the Sustainable Development Goals (SDGs). SDG 10– Reducing Inequalities—committed member states to reducing inequalities by promoting the inclusion of all population groups in socioeconomic and political spheres by 2030 [8]. Despite the increasing global focus on inequalities, current trajectories show that the world is unlikely to meet even 10% of the full targets under SDG 10 by 2030 [9]. People residing in the Global South, particularly in sub-Saharan Africa, bear the impact of the failure to achieve SDG10 [9].

Social protection programs can accelerate progress toward achieving SDG 10. Social protection is defined as policies and programs that help individuals and societies manage risk and uncertainty, protect them from poverty and inequality, and allow them to access economic opportunity [10]. They reduce poverty, inequality, and the prevalence of ill health; foster gender equality; and stimulate economic growth [11, 12, 13, 14]. Social protection programs that cater to the poorest populations can alleviate inequality [15]. Such programs are pro-poor. They prioritize the most impoverished and vulnerable people, including children, women, persons with disabilities, and the elderly [11, 16, 17, 18]. However, research investigating socioeconomic inequalities in receiving social protection in sub-Saharan Africa is limited. We assessed socioeconomic inequalities in receiving social protection among the general population, people (women and men) living with HIV (PLHIV), and adolescent girls and young women (AGYW) using population-based impact assessment survey data from 13 sub-Saharan African countries. Our hypothesis was that social protection was pro-poor, focused on people from poor households who are considered most vulnerable and deserving of access to it.

## 2 Materials and Methods

We analyzed SDG Indicator 1.3.1, defined as the proportion of the population receiving at least one social protection benefit from any source, as the main outcome indicator. UNAIDS earmarked this indicator as a target to measure the coverage of social protection for people living with, at risk of, or affected by HIV [19]. The target states ensure that by 2025, 45% of people living with, at risk of or affected by HIV have access to social protection benefits [19]. We examined inequalities in receiving social protection within the preceding 12 months of the survey interview by the general population, men and women living with HIV (MLHIV and WLHIV), and AGYW. We also assessed whether social protection programs in the participating countries reached the poorest households measured based on living standards including household assets.

We analyzed the PHIA survey data for countries with data on social protection receipt by the general population, MLHIV and WLHIV, and AGYW. The PHIA surveys collected a range of health and sociodemographic data to evaluate the impact of HIV programs in the countries supported by the United States President’s Emergency Plan for AIDS Relief. We used the Household, Adult Interview, and Adult HIV Biomarker datasets. In participating households, a household questionnaire was administered to the household head, who indicated all individuals living in the household and provided information on the household, such as assets, living standards, and access to social protection benefits. Individual questionnaires were then administered to eligible and consenting adults aged 15 or older in the household. The Adult HIV Biomarker data set contained the HIV test results of all adults and adolescents aged 15 or older who completed an individual interview and consented or agreed to provide blood samples for HIV testing. The interviews assessed wealth, education level, and other sociodemographic characteristics at the individual and household levels. They also included questions about external economic support. In addition, the questions identified AGYWs aged 15–24 years. We obtained the PHIA data sets from the PHIA Project website at https://phia-data.icap.columbia.edu/. We also conducted a desk review of social protection in the studied countries where information was available on the characteristics of social programs and their access by the general population, PLHIV, and AGYW.

### 2.1 Variables and outcome descriptions

Our primary outcome was the receipt of any form of external economic support. This support was defined as any external economic assistance provided to the household within the previous 12 months of the survey. This was derived from the PHIA household survey question “Has your household received any of the following forms of economic support in the last 12 months?” The acknowledgment of any economic support, including assistance for school fees, material support for education, food assistance, support for income generation, social pensions, and cash transfers, including pensions, disability, and child grants, was recorded as receiving social protection benefits. We classified a respondent as HIV-positive if the respondent self-reported an HIV-positive test result and the result of their HIV biomarker test was positive. AGYW were defined as females aged 15–24 years, men as males 15 years and older and women as females 15 years and older. Other explanatory variables included HIV prevalence, age, sex, marital status, household size, residence, employment, education, wealth, and region defined in Supplementary Table 1.

The study sample included women and men aged 15–59 years who were interviewed. Any individual with a missing sex was excluded from the analysis. Household wealth was evaluated via a composite measure reflecting living standards, based on asset ownership, which included items such as television sets, refrigerators, water access, and roofing. Region reflected the subregion of a country and was included in the analyses to account for subregional variation in access to social protection.

### 2.2 Analysis

#### 2.2.1 Measuring the level of social protection coverage

We used the *surveymeans* procedure to determine the weighted proportion of persons who reported receiving any social protection benefit for each country and population group – the public aged 15 to 59, MLHIV, WLHIV and AGYW. Survey weights accounting for nonresponse using Chi-squared automatic interaction detector analysis, noncoverage, and the probability of selection were applied. We used individual interview weights in the analysis of the data. Variances and 95% confidence intervals (CIs) were estimated using the corresponding jackknife replicate weights [20].

#### 2.2.2 Measuring inequality in social protection coverage

We used two methodologies to examine income-related inequality in receiving social protection. First, we constructed concentration curves for receiving social protection for each subpopulation within each country. A concentration curve represents the cumulative percentage of a variable of interest (social protection coverage in the context of this study) plotted on the y-axis against the cumulative proportion of the population—ranked by socioeconomic status from the poorest to the richest—on the x-axis [21]. The concentration curve coincides with the 45° line, known as the line of equality, when every individual receives the same value of the variable of interest. A concentration curve lying above (below) the line of equality signifies that the variable of interest is concentrated among the poor (rich). The degree of pro-poor (pro-rich) inequality increases as the curve diverges further above (below) the line of equality. In this study, we defined pro-poor social protection by the concentration curve of receiving social protection above the line of equality.

In the second approach, we computed the Concentration Index (CIX). The CIX encapsulates the information conveyed by the concentration curve, quantifying the socioeconomic inequalities associated with the variable of interest. The CIX is twice the area between the concentration curve and the line of equality, equating to zero in the absence of economic-related inequality [21]. A negative (positive) CIX value signifies that the curve lies above (below) the line of equality, indicating a disproportionate concentration of the variable of interest among the poor (rich). A zero CIX value can also occur if the curve intersects the line of equality and the areas above and below the equality line offset each other. In standard practice, CIX is interpreted in conjunction with the concentration curve. We conducted all analyses using Stata version 18.

#### 2.2.3 Ethics Statement

PHIA survey administration follows international scientific research standards in human subjects, including protecting respondents’ privacy and confidentiality of information. Each country’s PHIA survey report provides details of the survey design, sampling procedure, protection of the privacy and confidentiality of information, and obtaining informed consent.

Ethics and regulatory bodies, including ministries of health and institutional review boards, approved the PHIA survey protocols, consent forms, questionnaires, and other survey documents in each country. The institutional review boards of Columbia University Medical Center, Westat, and the Centers for Disease Control also reviewed and approved the survey documents.

This study did not require ethical clearance because the data were de-identified. It can be accessed by registering at the PHIA Project website at https://phia-data.icap.columbia.edu/

## 3. Results

The 13 countries and the years of the surveys were Cameroon (2017–2018), Côte d’Ivoire (2017– 2018), Ethiopia (2017–2018), Eswatini (2016–2017), Kenya (2018–2019), Lesotho (2016–2017), Malawi (2015–2016), Namibia (2017), Rwanda (2018), Tanzania (2016–2017), Uganda (2016–2017), Zambia (2016), and Zimbabwe (2015–2016). The sample size ranged from 10,197 in Eswatini to 29,577 in Tanzania, with median ages ranging between 27 years (interquartile range, IQR, 20–37 in Uganda) to 32 years (IQR 25–41 in. Kenya) (Table 1). HIV prevalence was lowest in Côte d’Ivoire (2.7%, 95% CI (2.4%–3.1%)) and highest in Eswatini (27.9%, 26.5%–29.3%). Women comprised 60% or more of people living with HIV in the surveyed countries (Supplementary Table 2).

**Table 1:** Survey weighted sample descriptive statistics by country (PHIA 2015-2019). The results are reported as (percentages with, sample size, 95% confidence intervals and absolute numbers unless otherwise indicated)

|  | **Cameroon**<br>**(N=26039)** | **Côte d'Ivoire**<br>**(N=18339)** | **Eswatini (N=10197)** | **Ethiopia (N=18466)** | **Kenya (N=23536)** | **Lesotho (N=12842)** | **Malawi (N=19092)** |
| --- | --- | --- | --- | --- | --- | --- | --- |
| Sex |  |  |  |  |  |  |  |
| Male | 49.1 (49.0 - 49.2) 11827 | 51.3 (51.2 - 51.4) 9145 | 45.5 (45.5 - 45.6) 4377 | 50.0 (49.9 - 50.1) 7735 | 47.5 (47.1 - 47.9) 9468 | 50.1 (50.0 - 50.1) 5339 | 48.5 (48.5 - 48.5) 8002 |
| Female | 50.9 (50.8 - 51.0) 14212 | 48.7 (48.6 - 48.8) 9194 | 54.5 (54.4 - 54.5) 5820 | 50.0 (49.9 - 50.1) 11731 | 52.5 (52.1 - 52.9) 14068 | 49.9 (49.9 - 50.0) 7503 | 51.5 (51.5 - 51.5) 11090 |
| Age [Years] (IQR) | 29 (21 - 39) | 29 (22 - 39) | 28 (21 - 38) | 28 (21 - 38) | 32 (25 - 41) | 30 (22 - 40) | 28 (20 - 38) |
| 15-24 | 36.6 (36.6 - 36.7) 9333 | 34.3 (34.2 - 34.4) 6199 | 37.2 (37.2 - 37.3) 3785 | 35.7 (35.6 - 35.8) 7882 | 21.9 (21.4 - 22.5) 4513 | 34.1 (34.0 - 34.2) 4403 | 39.8 (39.8 - 39.9) 7166 |
| 25-34 | 28.1 (28.0 - 28.2) 7474 | 31.2 (31.2 - 31.3) 5387 | 29.0 (29.0 - 29.1) 2843 | 31.3 (31.2 - 31.3) 5982 | 35.0 (34.7 - 35.2) 7857 | 29.9 (29.8 - 29.9) 3640 | 27.6 (27.6 - 27.6) 5489 |
| 35-44 | 19.4 (19.4 - 19.5) 4928 | 19.2 (19.2 - 19.3) 3818 | 18.7 (18.6 - 18.7) 1820 | 19.4 (19.3 - 19.4) 3288 | 23.8 (23.5 - 24.0) 5770 | 19.0 (18.9 - 19.0) 2387 | 18.2 (18.2 - 18.2) 3690 |
| 45-54 | 11.8 (11.7 - 11.8) 3084 | 11.6 (11.6 - 11.6) 2221 | 11.3 (11.3 - 11.3) 1256 | 10.4 (10.3 - 10.4) 1730 | 14.5 (14.4 - 14.7) 3911 | 12.1 (12.0 - 12.1) 1593 | 10.8 (10.8 - 10.8) 2050 |
| 55+ | 4.0 (4.0 - 4.1) 1220 | 3.6 (3.6 - 3.6) 714 | 3.8 (3.8 - 3.9) 493 | 3.3 (3.3 - 3.4) 584 | 4.8 (4.8 - 4.9) 1485 | 5.0 (4.9 - 5.0) 819 | 3.6 (3.6 - 3.6) 697 |
| Residence type |  |  |  |  |  |  |  |
| Rural | 47.4 (43.4 - 51.4) 14731 | 37.4 (33.8 - 41.2) 8762 | 72.0 (69.7 - 74.2) 7835 |  | 60.5 (58.5 - 62.5) 14548 | 58.8 (56.4 - 61.2) 7774 | 79.9 (77.0 - 82.5) 11827 |
| Urban | 52.6 (48.6 - 56.6) 11308 | 62.6 (58.8 - 66.2) 9577 | 28.0 (25.8 - 30.3) 2362 | 100.0 (89.9 - 100.1) 18466 | 39.5 (37.5 - 41.5) 8988 | 41.2 (38.8 - 43.6) 5068 | 20.1 (17.5 - 23.0) 7265 |
| Marital status |  |  |  |  |  |  |  |
| Single | 17.1 (16.4 - 17.7) 4402 | 8.8 (8.0 - 9.6) 1482 | 12.2 (11.5 - 13.0) 1290 | 14.8 (14.1 - 15.5) 3124 | 14.7 (14.0 - 15.3) 3670 | 17.0 (16.3 - 17.7) 2371 | 12.0 (11.3 - 12.7) 2441 |
| Married | 82.9 (82.3 - 83.6) 21637 | 91.2 (90.4 - 92.0) 16857 | 87.8 (87.0 - 88.5) 8907 | 85.2 (84.5 - 85.9) 16342 | 85.3 (84.7 - 86.0) 19866 | 83.0 (82.3 - 83.7) 10471 | 88.0 (87.3 - 88.7) 16651 |
| Household size (n) |  |  |  |  |  |  |  |
| 1 to 3 | 23.2 (21.9 - 24.6) 5823 | 25.7 (24.2 - 27.2) 4752 | 31.2 (29.2 - 33.3) 3146 | 39.5 (38.0 - 41.1) 8075 | 32.0 (30.5 - 33.5) 6774 | 40.5 (39.1 - 42.0) 5259 | 25.2 (23.9 - 26.5) 5075 |
| 4 to 6 | 35.8 (34.4 - 37.3) 9022 | 34.8 (32.6 - 37.0) 6308 | 37.4 (35.7 - 39.1) 3795 | 45.0 (43.7 - 46.3) 8361 | 44.8 (43.5 - 46.0) 10649 | 43.3 (41.8 - 44.8) 5543 | 50.8 (49.3 - 52.2) 9597 |
| >7 | 40.9 (38.9 - 43.1) 11194 | 39.6 (37.0 - 42.2) 7279 | 31.4 (29.2 - 33.7) 3256 | 15.5 (14.0 - 17.0) 3030 | 23.2 (22.0 - 24.6) 6113 | 16.1 (14.8 - 17.6) 2040 | 24.0 (22.6 - 25.5) 4420 |
| Employment status |  |  |  |  |  |  |  |
| Not employed | 45.1 (43.8 - 46.5) 12218 | 52.7 (51.1 - 54.3) 9962 | 56.7 (55.2 - 58.2) 5975 | 52.2 (50.8 - 53.7) 10996 | 42.4 (41.1 - 43.8) 11382 | 60.9 (59.6 - 62.1) 8156 | 70.4 (69.3 - 71.5) 13156 |
| Employed | 54.9 (53.5 - 56.2) 13821 | 47.3 (45.7 - 48.9) 8377 | 43.3 (41.8 - 44.8) 4222 | 47.8 (46.3 - 49.2) 8470 | 57.6 (56.2 - 58.9) 12154 | 39.1 (37.9 - 40.4) 4686 | 29.6 (28.5 - 30.7) 5936 |
| Education level |  |  |  |  |  |  |  |
| Not educated | 13.0 (11.9 - 14.3) 4695 | 40.8 (38.5 - 43.1) 7984 | 3.5 (3.0 - 4.0) 384 | 11.3 (10.3 - 12.4) 2318 | 7.5 (6.7 - 8.3) 2650 | 5.0 (4.5 - 5.5) 620 | 8.6 (8.0 - 9.3) 1521 |
| Primary | 26.2 (24.9 - 27.5) 7358 | 25.6 (24.3 - 26.9) 4904 | 25.9 (24.7 - 27.3) 2795 | 35.1 (33.7 - 36.6) 6821 | 49.5 (48.2 - 50.8) 12002 | 39.8 (38.2 - 41.4) 5247 | 63.2 (61.7 - 64.7) 10894 |
| Secondary | 33.5 (32.5 - 34.6) 8146 | 26.9 (25.4 - 28.5) 4539 | 59.9 (58.4 - 61.3) 6032 | 29.1 (28.0 - 30.2) 5706 | 30.9 (29.7 - 32.1) 6341 | 44.6 (43.2 - 46) 5719 | 25.3 (23.9 - 26.7) 5689 |
| Higher | 27.3 (25.4 - 29.3) 5840 | 6.7 (5.5 - 8.2) 912 | 10.7 (9.4 - 12.2) 986 | 24.5 (22.7 - 26.3) 4621 | 12.1 (11.2 - 13.1) 2543 | 10.6 (9.6 - 11.7) 1256 | 2.9 (2.6 - 3.3) 988 |
| Wealth quintiles: |  |  |  |  |  |  |  |
| Q1: Poorest | 19.9 (17.7 - 22.4) 7529 | 25.4 (22.0 - 29.1) 2867 | 20.2 (18.5 - 22.1) 2224 | 16.5 (14.4 - 18.9) 3379 | 19.9 (18.4 - 21.5) 6151 | 17.3 (15.5 - 19.3) 2485 | 15.2 (14.0 - 16.5) 2228 |
| Q2 | 20.1 (17.8 - 22.7) 5776 | 19.1 (16.5 - 22.0) 3304 | 19.9 (18.2 - 21.7) 2139 | 17.6 (16.3 - 19.0) 3495 | 20.5 (19.3 - 21.8) 4937 | 18.9 (17.6 - 20.4) 2530 | 18.2 (16.9 - 19.5) 2725 |
| Q3 | 21.3 (19.6 - 23.0) 4864 | 18.4 (16.4 - 20.6) 4169 | 22.9 (20.8 - 25.1) 2374 | 19.8 (18.4 - 21.3) 3895 | 20.7 (19.5 - 21.8) 4867 | 20.1 (18.6 - 21.7) 2539 | 20.1 (18.9 - 21.4) 3040 |
| Q4 | 18.9 (17.2 - 20.7) 4065 | 19.7 (17.8 - 21.7) 4508 | 18.0 (16.1 - 20.1) 1697 | 22.1 (20.2 - 24.0) 4212 | 20.2 (18.6 - 21.9) 4441 | 21.0 (19.6 - 22.5) 2592 | 22.1 (20.7 - 23.7) 3902 |
| Q5: Wealthiest | 19.8 (17.2 - 22.6) 3805 | 17.3 (14.5 - 20.6) 3491 | 19.0 (16.2 - 22.2) 1763 | 24.1 (21.8 - 26.5) 4485 | 18.8 (17.1 - 20.6) 3140 | 22.6 (20.7 - 24.8) 2696 | 24.4 (22.3 - 26.5) 7197 |

**Table 1: Survey weighted sample descriptive statistics by country (PHIA 2015-2019). The results are reported as percentages with 95% confidence intervals and absolute numbers unless otherwise indicated).**
|  | Namibia (N=18009) | Rwanda (N=29510) | Tanzania (N=29577) | Uganda (N=28212) | Zambia (N=21138) | Zimbabwe (N=21424) |
| --- | --- | --- | --- | --- | --- | --- |
| Sex |  |  |  |  |  |  |
| Male | 48.3 (48.2 - 48.3) 7967 | 48.1 (48.1 - 48.1) 13299 | 49.1 (49.1 - 49.1) 12867 | 47.4 (47.3 - 47.4) 12004 | 48.9 (48.9 - 49.0) 9104 | 47.7 (47.6 - 47.7) 8831 |
| Female | 51.7 (51.7 - 51.8) 10042 | 51.9 (51.9 - 51.9) 16211 | 50.9 (50.9 - 50.9) 16710 | 52.6 (52.6 - 52.7) 16208 | 51.1 (51.0 - 51.1) 12034 | 52.3 (52.3 - 52.4) 12593 |
| Age [Years] (IQR) | 29 (21 - 40) | 29 (21 - 39) | 28 (20 - 39) | 27 (20 - 37) | 27 (20 - 38) | 29 (21 - 38) |
| 15-24 | 35.2 (35.1 - 35.2) 6081 | 35.7 (35.7 - 35.7) 11365 | 38.4 (38.4 - 38.5) 10704 | 43.4 (43.3 - 43.4) 11321 | 41.1 (41.0 - 41.2) 8043 | 37.8 (37.8 - 37.8) 7730 |
| 25-34 | 28.7 (28.6 - 28.7) 4871 | 28.9 (28.8 - 28.9) 8181 | 27.3 (27.3 - 27.4) 8127 | 26.8 (26.8 - 26.9) 7643 | 27.3 (27.2 - 27.3) 5736 | 28.4 (28.4 - 28.4) 5570 |
| 35-44 | 19.4 (19.3 - 19.4) 3657 | 19.5 (19.5 - 19.5) 5636 | 18.8 (18.8 - 18.8) 5875 | 16.5 (16.4 - 16.5) 4923 | 18.1 (18.1 - 18.2) 4146 | 19.6 (19.6 - 19.6) 4360 |
| 45-54 | 12.5 (12.5 - 12.6) 2450 | 11.1 (11.1 - 11.1) 3086 | 11.5 (11.5 - 11.5) 3671 | 10.1 (10.1 - 10.1) 3315 | 10.1 (10.1 - 10.2) 2418 | 10.1 (10.1 - 10.1) 2596 |
| 55+ | 4.3 (4.3 - 4.3) 950 | 4.8 (4.8 - 4.8) 1242 | 3.9 (3.9 - 4.0) 1200 | 3.3 (3.2 - 3.3) 1010 | 3.4 (3.3 - 3.4) 795 | 4.1 (4.1 - 4.2) 1168 |
| Residence |  |  |  |  |  |  |
| Rural | 41.7 (39.4 - 44.0) 10146 | 79.3 (75.2 - 82.9) 21969 | 62.5 (58.7 - 66.2) 19601 | 71.3 (67.3 - 74.9) 20515 | 54.3 (50.8 - 57.6) 11911 | 64.0 (62.3 - 65.6) 14924 |
| Urban | 58.3 (56.0 - 60.6) 7863 | 20.7 (17.1 - 24.8) 7541 | 37.5 (33.8 - 41.3) 9976 | 28.7 (25.1 - 32.7) 7697 | 45.7 (42.4 - 49.2) 9227 | 36.0 (34.4 - 37.7) 6500 |
| Marital status |  |  |  |  |  |  |
| Single | 18.7 (17.8 - 19.6) 3467 | 11.0 (10.6 - 11.5) 3250 | 13.4 (12.8 - 14.0) 4085 | 16.9 (16.3 - 17.5) 4866 | 11.4 (10.9 - 11.9) 2612 | 13.0 (12.4 - 13.6) 3111 |
| Married | 81.3 (80.4 - 82.2) 14542 | 89.0 (88.5 - 89.4) 26260 | 86.6 (86.0 - 87.2) 25492 | 83.1 (82.5 - 83.7) 23346 | 88.6 (88.1 - 89.1) 18526 | 87.0 (86.4 - 87.6) 18313 |
| Household size (n) |  |  |  |  |  |  |
| 1 to 3 | 30.9 (29.5 - 32.4) 5294 | 15.2 (14.3 - 16.2) 4595 | 24.0 (22.7 - 25.4) 6749 | 19.5 (18.6 - 20.5) 5018 | 18.1 (17.1 - 19.2) 3836 | 27.8 (26.6 - 29.1) 6035 |
| 4 to 6 | 33.7 (32.1 - 35.4) 5783 | 44.3 (42.8 - 45.8) 12971 | 41.1 (39.8 - 42.4) 11799 | 35.7 (34.5 - 37.0) 9730 | 42.9 (41.8 - 44.1) 9176 | 49.4 (48.0 - 50.8) 10403 |
| >7 | 35.4 (33.5 - 37.2) 6932 | 40.5 (38.6 - 42.4) 11944 | 34.9 (33.1 - 36.7) 11029 | 44.8 (43.2 - 46.3) 3464 | 38.9 (37.5 - 40.4) 8126 | 22.8 (21.4 - 24.2) 4986 |
| Employment status |  |  |  |  |  |  |
| Not employed | 54 (52.8 - 55.3) 10329 | 59.5 (58.3 - 60.6) 17441 | 55.3 (54.2 - 56.4) 16829 | 47.0 (45.9 - 48.0) 13776 | 66.2 (65.0 - 67.4) 14295 | 59.8 (58.6 - 60.9) 13423 |
| Employed | 46.0 (44.7 - 47.2) 7680 | 40.5 (39.4 - 41.7) 12069 | 44.7 (43.6 - 45.8) 12748 | 53.0 (52.0 - 54.1) 14436 | 33.8 (32.6 - 35.0) 6843 | 40.2 (39.1 - 41.4) 8001 |
| Education level |  |  |  |  |  |  |
| Not educated | 6.7 (6.0 - 7.4) 1579 | 9.1 (8.6 - 9.6) 2502 | 12.9 (11.9 - 14.0) 4403 | 7.1 (6.6 - 7.6) 2486 | 4.9 (4.3 - 5.7) 1121 | 1.9 (1.7 - 2.1) 518 |
| Primary | 23.3 (22.2 - 24.5) 4994 | 61.6 (60.3 - 62.8) 17643 | 61.5 (60.4 - 62.6) 18231 | 55.7 (54.3 - 57.0) 16061 | 41.9 (40.2 - 43.6) 9164 | 25.1 (24.1 - 26.2) 6143 |
| Secondary | 58.0 (56.4 - 59.5) 9972 | 25.3 (24.2 - 26.3) 7921 | 20.3 (19.3 - 21.4) 5593 | 26.2 (25.3 - 27.2) 6819 | 44.8 (43.2 - 46.3) 9170 | 64.8 (63.6 - 66.0) 13314 |
| Higher | 12.0 (10.5 - 13.6) 1464 | 4.1 (3.6 - 4.7) 1444 | 5.3 (4.7 - 5.9) 1350 | 11.1 (10.2 - 12.0) 2846 | 8.4 (7.4 - 9.6) 1683 | 8.2 (7.2 - 9.3) 1449 |
| Wealth quintiles |  |  |  |  |  |  |
| Q1: Poorest | 19.2 (17.8 - 20.7) 4610 | 18.7 (16.8 - 20.7) 5054 | 18.6 (16.4 - 21.1) 6155 | 20.5 (19.2 - 22.0) 7631 | 15.0 (13.6 - 16.5) 3357 | 19.2 (17.7 - 20.9) 4965 |
| Q2 | 19.8 (18.0 - 21.8) 4122 | 19.0 (17.5 - 20.6) 5275 | 20.3 (18.7 - 21.9) 6154 | 19.6 (18.1 - 21.2) 5618 | 18.0 (16.6 - 19.5) 3931 | 19.6 (18.5 - 20.8) 4487 |
| Q3 | 21.4 (19.4 - 23.5) 3827 | 19.9 (18.6 - 21.2) 5543 | 20.8 (19.3 - 22.4) 6583 | 19.7 (18.3 - 21.2) 5240 | 20.0 (18.3 - 21.8) 4274 | 19.2 (17.8 - 20.7) 4122 |
| Q4 | 20.3 (18.2 - 22.6) 3077 | 20.8 (19.3 - 22.3) 5856 | 19.6 (17.8 - 21.6) 5510 | 19.8 (18.4 - 21.3) 4611 | 21.7 (19.6 - 24.0) 4526 | 19.7 (17.8 - 21.9) 3637 |
| Q5: Wealthiest | 19.3 (16.7 - 22.1) 2373 | 21.7 (19.3 - 24.4) 7782 | 20.7 (18.7 - 22.8) 5175 | 20.3 (18.1 - 22.7) 5112 | 25.2 (22.7 - 28.0) 5050 | 22.2 (20.1 - 24.4) 4213 |

Q1 Quintiles 1: Poorest, Q5: Wealthiest. IQR Interquartile range

More than 60% of the respondents lived in rural areas in eight countries (i.e., Eswatini, Kenya, Lesotho, Malawi, Rwanda, Tanzania, Uganda, and Zimbabwe). In all countries surveyed, 80% or more of respondents were married or cohabiting, and 60% or more had at least four members. Of the respondents, 50%–70% were unemployed, except in Cameroon, Kenya, and Uganda, where less than 50% were unemployed. Up to 13% of the respondents had no formal education, except in Côte d’Ivoire, where 42% had no formal education. In general, the proportion of respondents from wealth quintile one (Q1), that is, the bottom 20% of households, was similar to those from Q2–Q5 (Table 1).

The proportion of the general population receiving social protection varied from 5.2% (95% CI 4.5%– 6.0%) in Ethiopia to 39.9% (37.0%–42.8%) in Eswatini. Among PLHIV households, the proportion receiving social protection varied from 6.9% (5.7%–8.4%) among MLHIV in Zambia to 45.0% (41.2– 49.0) among WLHIV in Namibia. Among AGYW, the proportion varied from 4.4% (3.6–5.3) in Ethiopia to 44.6% (40.8–48.5) in Eswatini.

The proportion of the general population reporting receiving social protection from the poorest wealth quintile (Q1) ranged from 8.1% (6.4%–10.2%) in Cameroon to 56.2% (51.5%–60.7%) in Eswatini. Among the wealthiest quintiles (Q5), the proportion ranged from 3.6% (2.6%–5.0%) in Ethiopia to 19.7% (16.25–23.8%) in Namibia (Table 2). In general, 15% or less of the respondents from Q1 reported receiving social protection in eight countries (i.e., Cameroon, Côte d’Ivoire, Ethiopia, Kenya, Malawi, Tanzania, Uganda, and Zambia), with 10% or less in three countries (Cameroon, Côte d’Ivoire, and Ethiopia); 15%–20% in Rwanda, 30% in Zimbabwe, 40% in Lesotho, and more than 50% in Eswatini and Namibia.

**Table 2:**
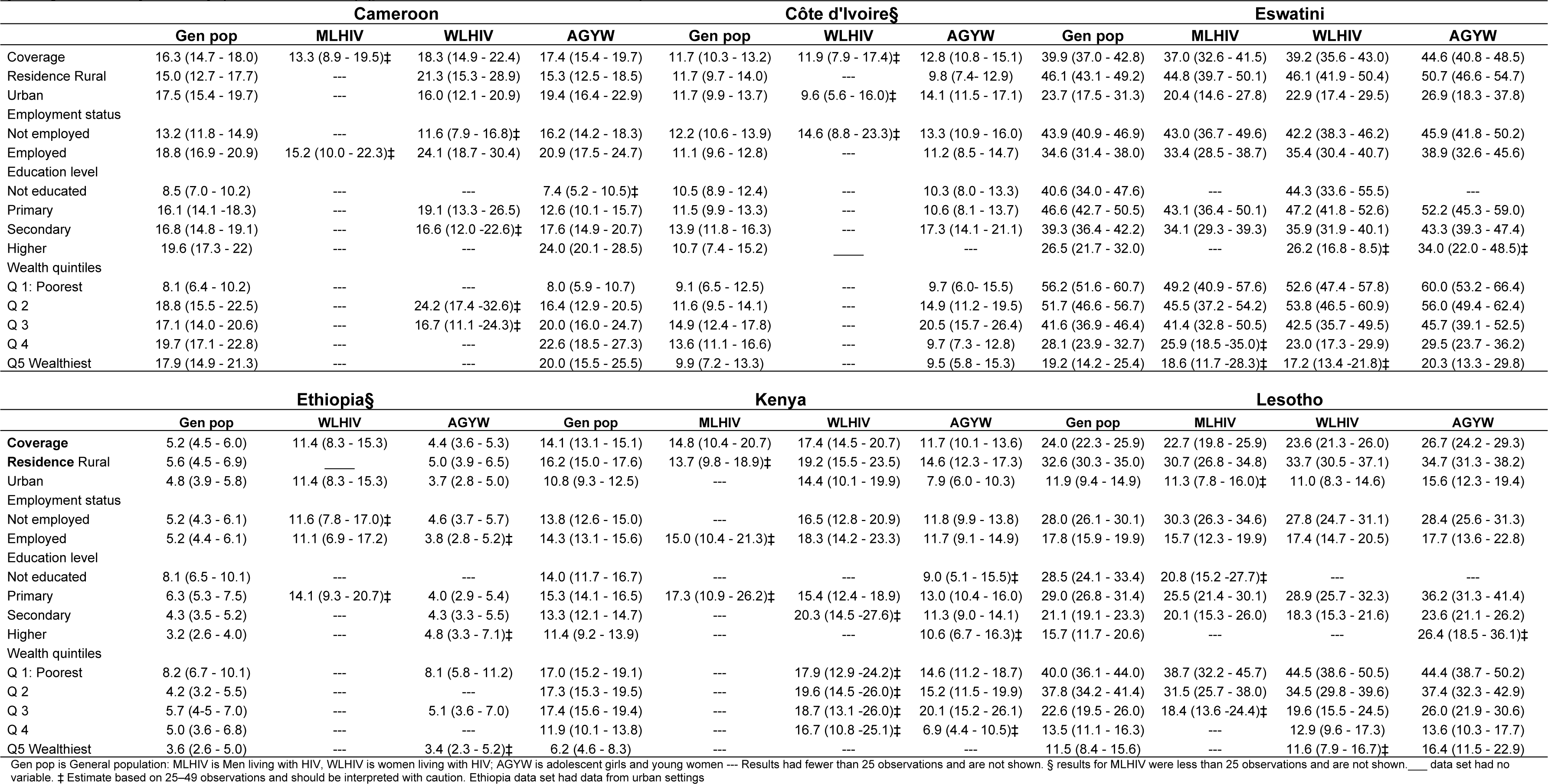
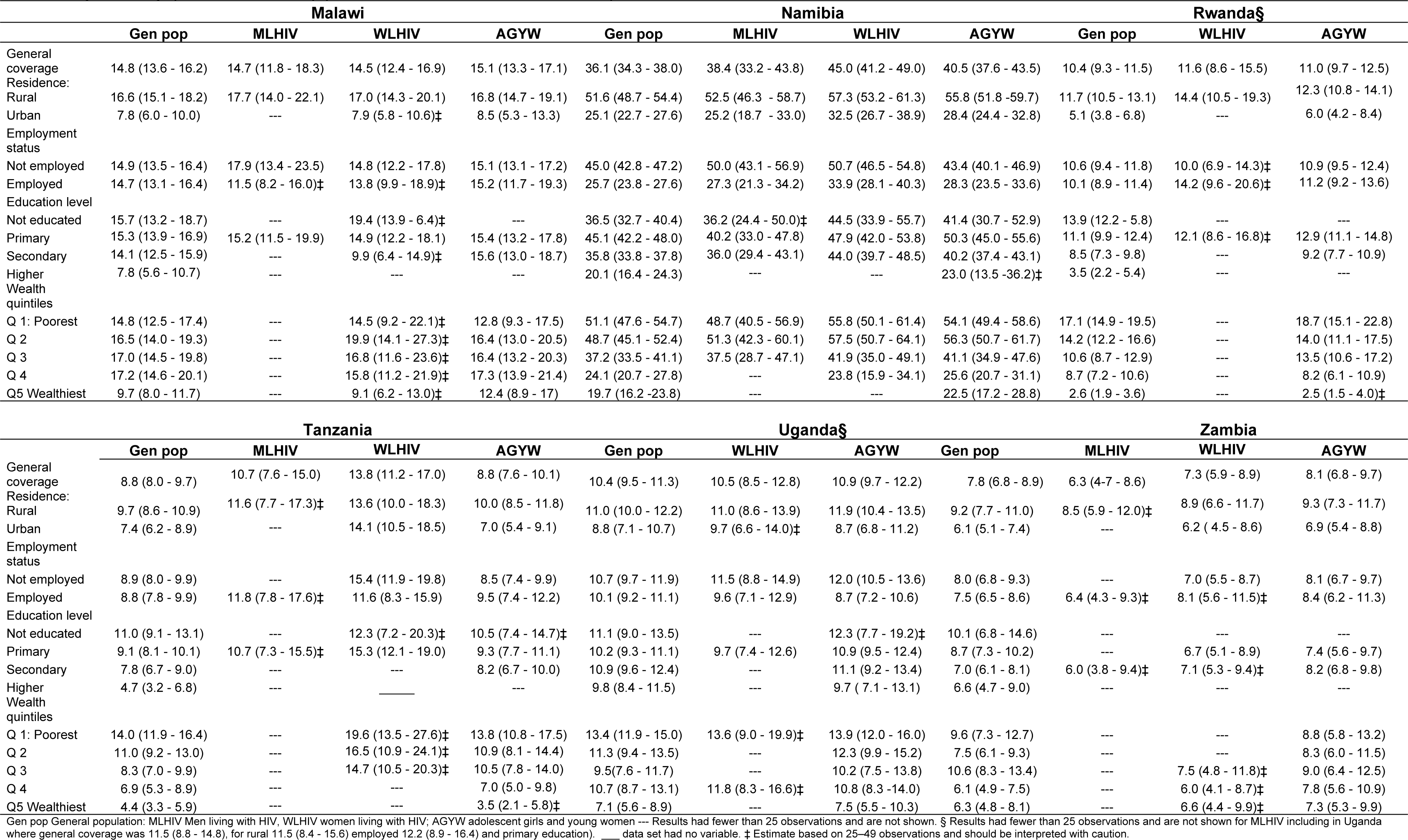
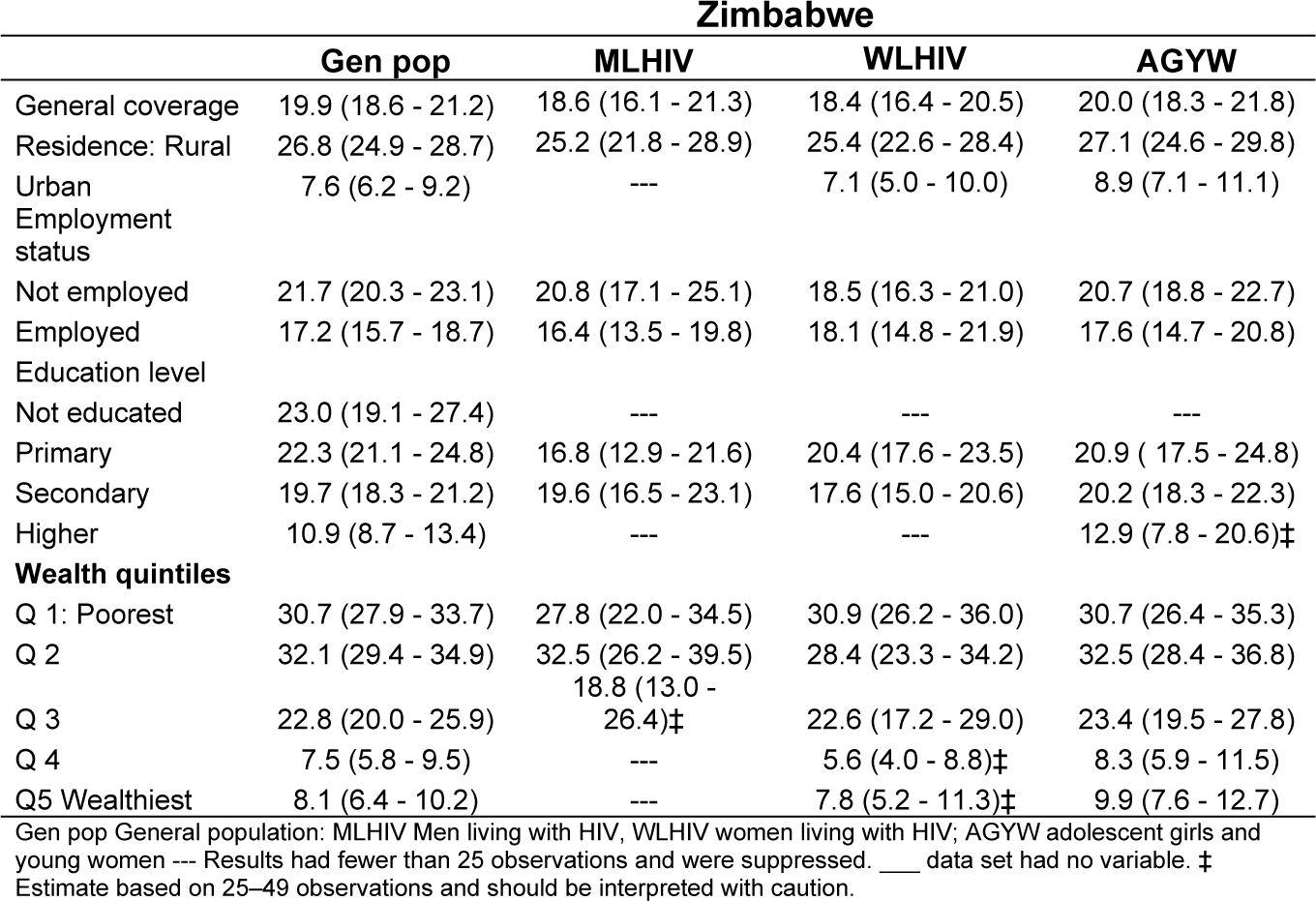
Survey weighted household social protection coverage for the general population, among people living with HIV (male and female), and adolescent girls and adolescent girls and young women by country (PHIA 2015-2019) (percent, 95% confidence interval)

Figure 1 presents the concentration curves of access to social protection by country for the general population, MLHIV and WLHIV, and AGYW. Table 3 reports the associated concentration indices. The results show that socioeconomic inequalities in access to social protection were pro-rich only in Cameroon, among the general population, and AGYW–evident as the concentration curves lie below the line of equality—with a CIX value of 0.122 (p < 0.001) among the general population and a CIX value of 0.169 (<0.001) among AGYW. This result shows that more people from wealthier than poor households reported receiving social protection. In Côte d’Ivoire, socioeconomic inequalities in receiving social protection were pro-rich, but the associated CIX estimates were not significantly different from zero. In the remaining 11 countries, social protection was pro-poor. The CIX values for socioeconomic inequalities in receiving social protection in these countries ranged from −0.080 (p = 0.002) among the general population in Malawi to −0.372 (p < 0.001) among WLHIV in Zimbabwe (Table 3).

**Figure 1:**
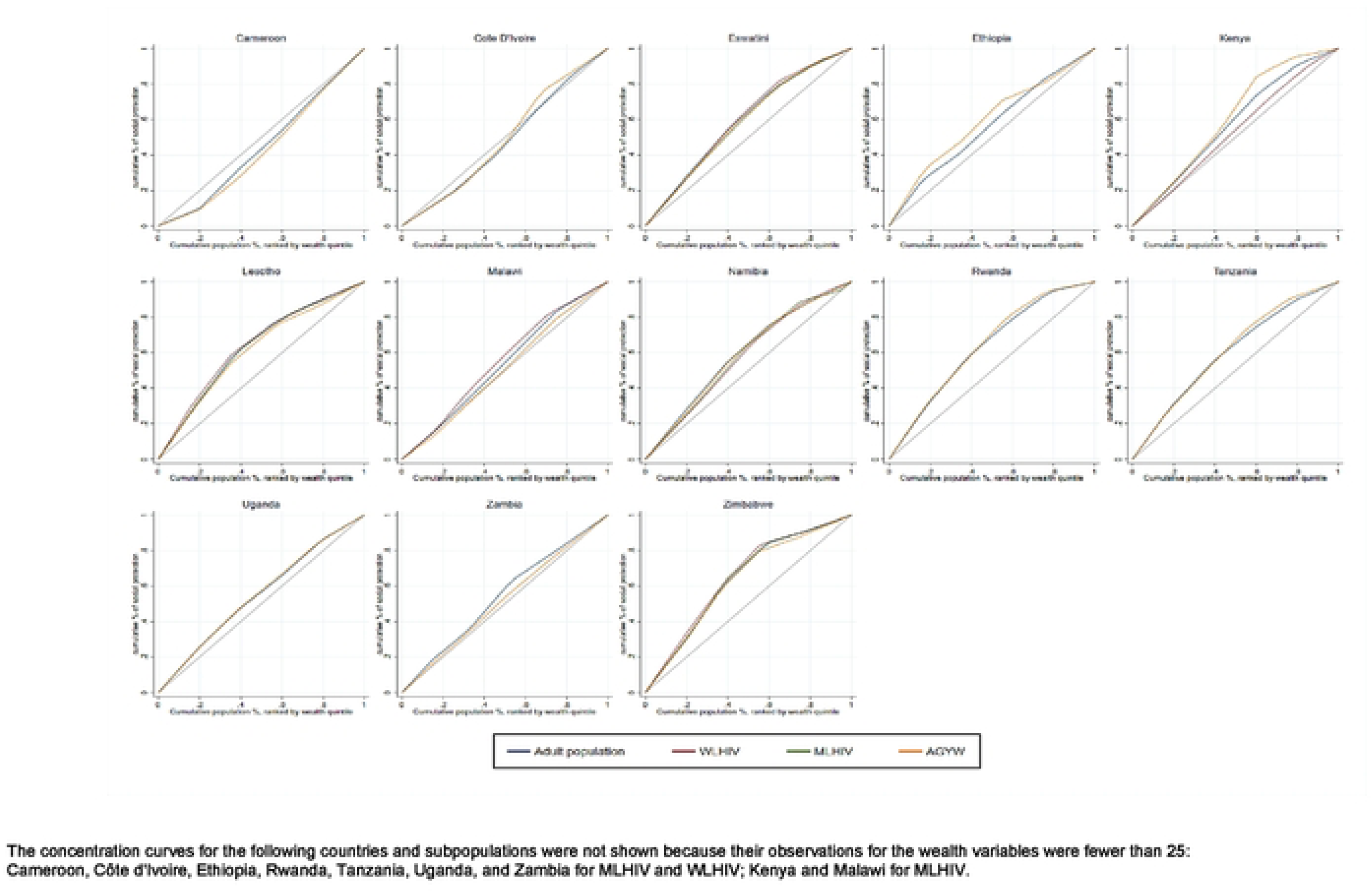
Concentration curves of receiving social protection among the general population, people living with HIV (women and men), and adolescent girls and young women in sub-Saharan African Countries (PHIA 2015-2019).

**Table 3:**
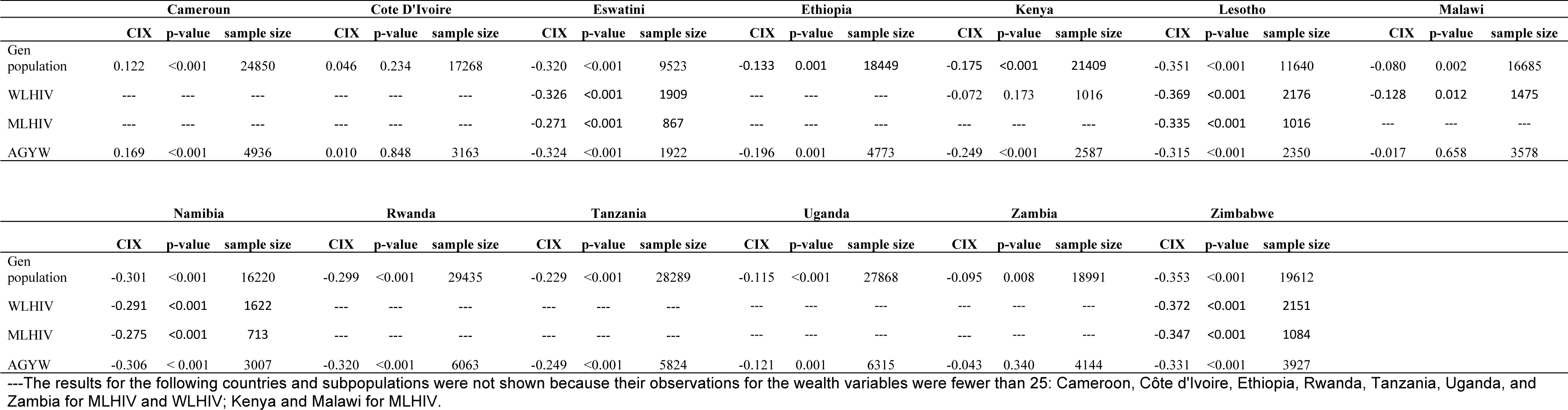
Survey weighted concentration index of socioeconomic inequalities in receiving social protection for the general population, people living with HIV (male and female) and adolescent girls and young women (CIX, p-value, sample.

In Eswatini, Lesotho, Rwanda, Namibia, and Zimbabwe, socioeconomic inequalities in receiving social protection were pro-poor, below −0.300, among all population groups. Socioeconomic inequalities in receiving social protection access were pro-poor and moderate, with CIX values ranging from −0.100 to −0.300 among the general population and AGYW in Ethiopia, Kenya, and Uganda and among the general population, WLHIV, and AGYW in Tanzania. Social protection was pro-poor and of low inequality, that is, between CIX −0.010 and CIX = −0.100 among the general population and MLHIV and WLHIV in Malawi, and the general population and MLHIV in Zambia (Figure 1 and Table 3).

## 4 Discussion

This study examined economic-related inequality in receiving social protection among the general population, MLHIV and WLHIV, and AGYW in 13 sub-Saharan African countries. The study also evaluated whether people in the poorest households received social protection. Our findings showed that the proportion of the general population receiving social protection varied from 5.2% (95% CI 4.5%–6.0%) in Ethiopia to 39.9% (37.0%–42.8%) in Eswatini. Social protection was pro-poor in 11 out of the 13 countries studied, implying that more people from poor households received social protection than those from wealthier households in these 11 countries. However, in eight of these 11 countries, less than 15% of people from the poorest quintile households reported receiving social protection. Cameroon was the only country where social protection was pro-rich. These results bear considerable policy implications for the targeting, scale up, and equalization of access to social protection among the general population, MLHIV and WLHIV, and AGYW.

Our study’s first results showed that the proportion of respondents receiving social protection varied widely, ranging from 4.4% among AGYW in Ethiopia to 44.6% among WLHIV in Namibia, corroborating the evidence. The International Labour Organisation (ILO) report 2020–2022 indicated that only 46.9% of the global population were covered by at least one social protection benefit in 2020 [22]. The ILO report further highlighted considerable regional disparities in access to social protection, with the lowest coverage in Africa at 17.4% and the highest in Europe and Central Asia at 83.9% [22]. A study covering Eswatini, Malawi, Tanzania, and Zambia found that the proportion receiving social protection varied from 7.7% in Zambia to 39.6% in Eswatini [23]. The paper found comparable social protection coverage among the AGYW and PLHIV to the general population in Malawi and Zambia [23].

Our second finding revealed that social protection was pro-poor in 11 of the 13 countries surveyed. The pro-poor social protection found in our study was expected and aligns with our hypothesis. Social protection programs are generally focused on the most impoverished households [11, 16, 17, 18]. In this regard, our findings are consistent with the core objectives of social protection, which prioritize the poorest households. However, in eight of the 11 countries, fewer than 15% of people from households in the bottom wealth quintile reported receiving social protection. A review of evidence from 123 countries found that only 22% of the poorest 20% received social assistance in support of our finding [24]. Cameroon stood out as an outlier, exhibiting pro-rich social protection, underscoring a significant shortfall in reaching people from the poorest households. A contributing factor to this disparity in Cameroon was the high access to social protection among employed individuals, indicating that the benefits were linked to their employment. For example, civil service pensions, which benefited only 141,000 pensioners in 2016 in Cameroon, were allocated over 10 times more funding by the government of Cameroon than all social assistance schemes combined [25]. Another potential reason was the relatively nascent state of social protection in Cameroon [25]. Despite the development of a comprehensive social protection policy in 2017, the program was not approved. Social protection programs remained small-scale and uncoordinated [25]. This result shows a potential gap in reaching people from the poorest households.

The limited coverage of individuals from the poorest quintile households identified in our study may be due to difficulties faced by low-income countries in identifying the poorest population groups to target their social protection services [24]. Another reason may be the dynamic mobility of people across economic groups. For example, in the Occupied Palestine State, where 40% of individuals receiving social protection were categorized as poor, they moved up and down income groups over time [26].

These findings underscore that pro-poor social protection alone is insufficient to reach people from the poorest households. Although policymakers may contemplate redistributing social protection benefits from wealthier to poor households, this approach may not be feasible or desirable. Wealthier households can descend into poverty, requiring social protection [26]. Another potential explanation could be the nontakeup of social protection benefits, a common phenomenon among marginalized populations who need social protection the most [27]. Nontakeup pertains to eligible individuals not accessing available benefits for a range of reasons, including lack of information, complex or costly procedures, limited access to digital technology and know-how, stigma, discrimination, shame, and fear of interacting with social services [27]. Moreover, people from the poorest households, eligible to access social protection, may not take up available social protection benefits owing to inadequate coverage and the narrow scope of programs [28]. According to our study, only in countries with an overall higher social protection coverage, such as Eswatini, Lesotho, Namibia, and Zimbabwe, the proportion of the poorest wealth quintile households reached were also high. A study examining global inequalities in accessing reproductive, maternal, newborn, and child health services showed that countries, with low inequality and high coverage in these services, effectively reached the poorest women and children [29]. The coverage of social protection needs to be broadened and deepened to reach the poorest households. Additionally, strategies to identify households that are thrust into poverty owing to emerging risks, such as financial crises, conflicts, droughts, disasters, and pandemics like COVID-19, and link them to social protection, should be developed.

This study has several limitations and strengths. Contrary to the ILO’s strategy of presenting summarized national responses to government-provided social protection [22], our study compiles individual responses from various countries through household surveys. Notwithstanding, our estimates correspond to the data from the ILO 2020–2022 report, indicating that our measurement reflects the same information that governments use in their reporting. Another limitation of our study is the absence of identification for marginalized people, such as gay men and other men who have sex with men, sex workers and migrants. Marginalized population groups suffer a bulk of hardships owing to inequalities [1, 2]. These population groups may be excluded from accessing social protection benefits often because of stigma, discrimination, and punitive laws [30]. The potential barriers to accessing social protection benefits of these subgroups were not addressed in this study. This gap stems from either a lack of available information or an insufficient sample size to conduct meaningful analysis. Furthermore, we did not disaggregate the specific social protection benefits received or their monetary value. Neither did we account for the rapid scale up of social protection in response to the COVID-19 pandemic, which would have provided more insights into our analysis.

## 5. Conclusion

In the countries surveyed, access to social protection for the general population, MLHIV and WLHIV, and AGYW was low but favored people from poor households. However, pro-poor social protection, although necessary, is not sufficient to ensure that people from the poorest households receive social protection. Further research is required to identify and reach people from the poorest households with social protection in sub-Saharan Africa.

## Data Availability

The data is publicly available and can be accessed upon request at the PHIA Project website at https://phia-data.icap.columbia.edu/

## Author contributions

DC designed the study. DC and SA-A collated the data. DC, SA-A, LH, ARH, MS, CM, JGJM, OK and JE performed the data analysis. DC and SA-A conducted the data visualization. DC and SA-A drafted and revised the manuscript. DC, SA-A, LH, ARH, MS, CM, JGJM, OK and JE critically reviewed the drafts. All authors approved the final manuscript.

## Acknowledgment

The authors acknowledge comments from UNAIDS.

## Disclaimer

Where authors are identified as personnel of the World Health Organization or UNAIDS, the authors alone are responsible for the views expressed in this article and they do not necessarily represent the decisions, policy, or views of the World Health Organization or UNAIDS.

## Supporting information

S1 Table. Variable descriptions.

S2 Table. Survey weighted proportions of men and women living with HIV by country (PHIA 2015-2019).

